# Attitudes towards the regulation and provision of abortion among healthcare professionals in Britain; cross sectional survey data from the SACHA study

**DOI:** 10.1101/2024.04.09.24305548

**Authors:** K Wellings, R Scott, S Sheldon, O McCarthy, P Palmer, R Meiksin, J Shawe, S Cameron, M Lewandowska, J Reiter, RS French, (and the SACHA study team #)

## Abstract

**Objectives:** Therapeutic, regulatory and technological changes have contributed to new directions in abortion care. We aimed to gather views of healthcare professionals on current and future regulation and provision of abortion in Britain.

**Design:** Cross sectional, stratified cluster sample survey with additional free text comments.

**Setting:** Healthcare settings in Britain

**Participants:** Healthcare professionals (nurses, doctors, midwives, pharmacists) working in a range of healthcare services, including dedicated abortion services.

**Measures:** Knowledge of and attitudes towards the regulation and provision of abortion.

**Results:** 771 healthcare professionals from all areas of Britain responded to the survey. Support for abortion being a woman’s choice was high, more than nine in ten saw it as such and a clear majority favoured abortion being treated as a health rather than a legal issue. Only 6.2% saw abortion at any gestational age as contrary to their personal beliefs and a similarly small minority (6.7%) were against abortion after 12 weeks gestation. One in five of all healthcare professionals and a third of those aged under 30 were unaware that the law in Britain requires two doctors to authorise an abortion. Free text comments revealed opposition to the need for this legal requirement. Support for an extended role for nurses in abortion care was high; two thirds (65.3%) agreed that nurses should be able to prescribe abortion medication. Little more than a third of all healthcare professionals (37%) agreed that abortion should be standard practice in their service; the proportion was highest among those in sexual and reproductive health services (58.4%) and lowest among those in general practice (18.7%).

**Conclusions:** Health care professionals in Britain are generally supportive of abortion being treated in the same way as other health issues and would be likely to support any moves to decriminalise abortion.

Strengths and limitations of the study

- The study describes the views of a range of healthcare professionals in different specialities and settings on the current and future regulation and provision of abortion.
- The use of a stratified cluster sampling strategy increases the generalisability of findings.
- Additional qualitative data from free text comments helps in understanding the survey responses.
- The main limitation stems from the timing of the study. Scheduling fieldwork at the height of the COVID-19 pandemic and its immediate aftermath may have introduced both participation and reporting bias.
- Healthcare professionals most actively involved in coping with the pandemic may have declined to participate because of time pressures and/or provided responses influenced by awareness of the current constraints of their workload.

What is already known on this topic

- The 21^st^ century has seen major changes in the landscape of abortion. Therapeutic and technological advances have led to an increase in the prevalence of medication abortion which can be safely managed by women at home and have provided the innovative telemedical interventions enabling them to do so.
- The trends have prompted reflection on the appropriate regulation of abortion in Britain, on the role of different cadres of healthcare professionals; and on the appropriate healthcare settings in which abortion can be provided. Little is known about the views of health care professionals on these issues.

What this study adds

- The law on abortion is not widely known. One in five of all health care professionals and one in three aged under 30 are not aware that for abortion to be legal in Britain two doctors must certify that certain grounds have been met.
- There is near universal support among healthcare professionals for abortion being a woman’s choice and a widely held view that abortion should be treated no differently from any other health condition.
- Enthusiasm for incorporating abortion into existing practice is high among healthcare professionals employed in sexual and reproductive health services but most of those in general practice see abortion provision as out of scope for their service.
- An extended role in abortion for nurses commands considerable support among healthcare professionals, including prescribing abortion medication and, whilst the legal requirement to do so remains, to authorise abortions.

How this study might affect research, practice or policy

- Should there be political will in Britain for a legal reform that removes specific criminal prohibitions against abortion, our data suggest that it would encounter strong support amongst healthcare professionals.

## Introduction

Recent years have seen marked changes in the landscape of abortion. Therapeutic advances have led to the increasing adoption of medication as opposed to surgical abortion^1^. Broader trends within 21^st^ century health systems have contributed to new directions in abortion provision: the increasing use of digital approaches in health care, task-sharing by healthcare professionals, and greater patient-centred care and supported self-management. Changes in the cultural climate have formed the backdrop to these trends, including increasing secularisation and heightened attention to reproductive rights and gender equality.

These developments have prompted re-examination of issues such as the appropriate location for procedures and the roles of healthcare cadres in abortion provision. They have also provided the impetus to changes in the regulation of abortion. Abortion in Britain continues to be regulated by the 1967 Abortion Act, which creates an exemption whereby abortion will not be deemed a criminal offence provided that certain conditions are met. Under the Act, for an abortion to be lawful, two doctors must sign certifying that specific grounds have been met, the abortion must be performed by a medical practitioner and must be carried out in an NHS hospital or other approved premises^2^. There have been significant changes in how the Act is implemented. The requirement for authorisation by two doctors remains, but in practice is liberally interpreted and may be based on information conveyed to clinicians by other health care staff^2.^. The requirement that abortions must be performed by a doctor is interpreted to permit other appropriately trained healthcare professionals to carry out certain tasks in abortion under the doctor’s supervision^2^. During the COVID-19 pandemic, the governments in England, Wales and Scotland approved home administration of mifepristone for early medication abortion with remote consultations and telemedical support. The change in the regulations was made permanent in April 2022 and the Abortion Act was amended accordingly in August 2022^3^.

In the recent past several countries have made liberalising changes to the legislative framework governing abortion.^4^ Among high income countries, abortion was decriminalised in New Zealand in 2020 and in all jurisdictions in Australia by 2022. In 2018-2019, the Belgian Parliament launched two legal initiatives challenging the role of criminal law in the regulation of abortion^5^. Decriminalisation of abortion in the Republic of Ireland in 2018, and in Northern Ireland and the Isle of Man in 2019^4^ led to speculation that Britain might follow^6^.

Pressure for the 1967 Abortion Act to be repealed in Britain has mounted, as evidenced in statements from the Royal Colleges and professional associations, in political party manifestos and in Private Member’s Bills in Britain ^7,8,9,10^. It echoes views expressed by international organisations such as the WHO and the Committee on the Elimination of Discrimination against Women (CEDAW) that abortion should not be criminalised ^11,12^.

Amending the legal framework for abortion is likely to create opportunities for a wider range of healthcare professionals to provide abortion care and support^13.^. Even within the existing legal framework, healthcare professionals from a range of specialities may encounter patients seeking abortion referral or support in routine health care given current prevalence. Nearly one in three women in Britain can now expect to experience abortion before age 45^14^ and demand is increasing 32^15^. Understanding the views of healthcare professionals is important to optimising service provision, yet little is known on the subject in Britain. Studies in the past have examined attitudes among selected groups such as medical students^16^, general practitioners (GPs)^17^ and obstetricians and gynaecologists^18^. There are no up-to-date, comprehensive data on the views of the wider range of healthcare professionals such as nurses, midwives and pharmacists.

In this paper we draw on data gathered as part of a larger NIHR-funded study: SACHA (Shaping Abortion for Change^2^), aimed at guiding the optimal configuration of health services in Britain in response to changes in models of abortion care. We report on attitudes towards the regulation and provision of abortion among a range of healthcare professionals.

## Methods

### Participants and procedures

We carried out a survey of knowledge, attitudes and experience of abortion provision between November 2021 and August 2022 in England, Scotland and Wales^19^. Healthcare professionals including nurses, midwives, doctors and pharmacists in a range of settings were eligible for inclusion. We used a stratified cluster sampling strategy, selecting a random sample of services as ‘clusters’. Service types were drawn from separate sampling frames. A list of general practices was compiled from data from the Care Quality Commission in England (CQC); Health Inspectorate Wales; and NHS Inform Scotland. A list of NHS and independent sector abortion services was compiled from abortion statistics in England and Wales reported to the Chief Medical Officer in 2020 and clinic lists available from the independent sector providers; and from communication with those involved in abortion provision in Scotland. Registered pharmacies in England, Scotland and Wales were identified via the General Pharmaceutical Council. For sexual and reproductive health, and midwifery services, our sampling frame consisted of a complete list of all six-digit postcodes in England, Wales, and Scotland. Randomly selected postcodes were entered into the ‘find a service’ function on the NHS website to identify the nearest service for selection. All eligible staff within each selected service were invited to take part. Practitioners within each service were identified from website staff profiles or, where not publicly available, by contacting service managers.

Prevalence estimates of pro-choice attitudes among healthcare professionals from earlier research in Britain^17^ informed sample size calculations. Following piloting for comprehension and time to completion, questionnaires, together with information sheets, consent forms, unconditional tokens of gratitude (vouchers) and free-post return envelopes, were mailed to individual practitioners within each service. The option of completing the survey online was provided. Non-responders were followed up with two reminder phone calls or emails. Unique ID numbers were provided to enable response rates to be calculated. The fully standardised survey questionnaire probed knowledge of and attitudes towards the regulation and provision of abortion care and support. Space was provided for free text comments to be added.

### Measures

Knowledge of the law was measured by probing awareness of the legal requirement for abortion to be signed off by a doctor. Views on regulatory aspects of abortion were measured by seeking agreement with statements relating to abortion as a woman’s choice; abortion as a health rather than a legal issue; gestational age limits; and the probity of abortion. Attitudes towards abortion provision were measured by seeking respondents’ views on whether abortion care should be standard practice in their specialty and on the ability of different practitioners to provide aspects of abortion care. Verbatim wording of statements is provided in Tables 1 and 2. Response options were three-point Likert scales (True, False and Don’t Know; and Agree; Neither Agree nor Disagree; Disagree). Respondent characteristics included in the analysis were gender, age, time since qualification, current involvement in abortion provision; service type, profession, constituent country of Britain and importance of religion and political persuasion.

**Table 1.**
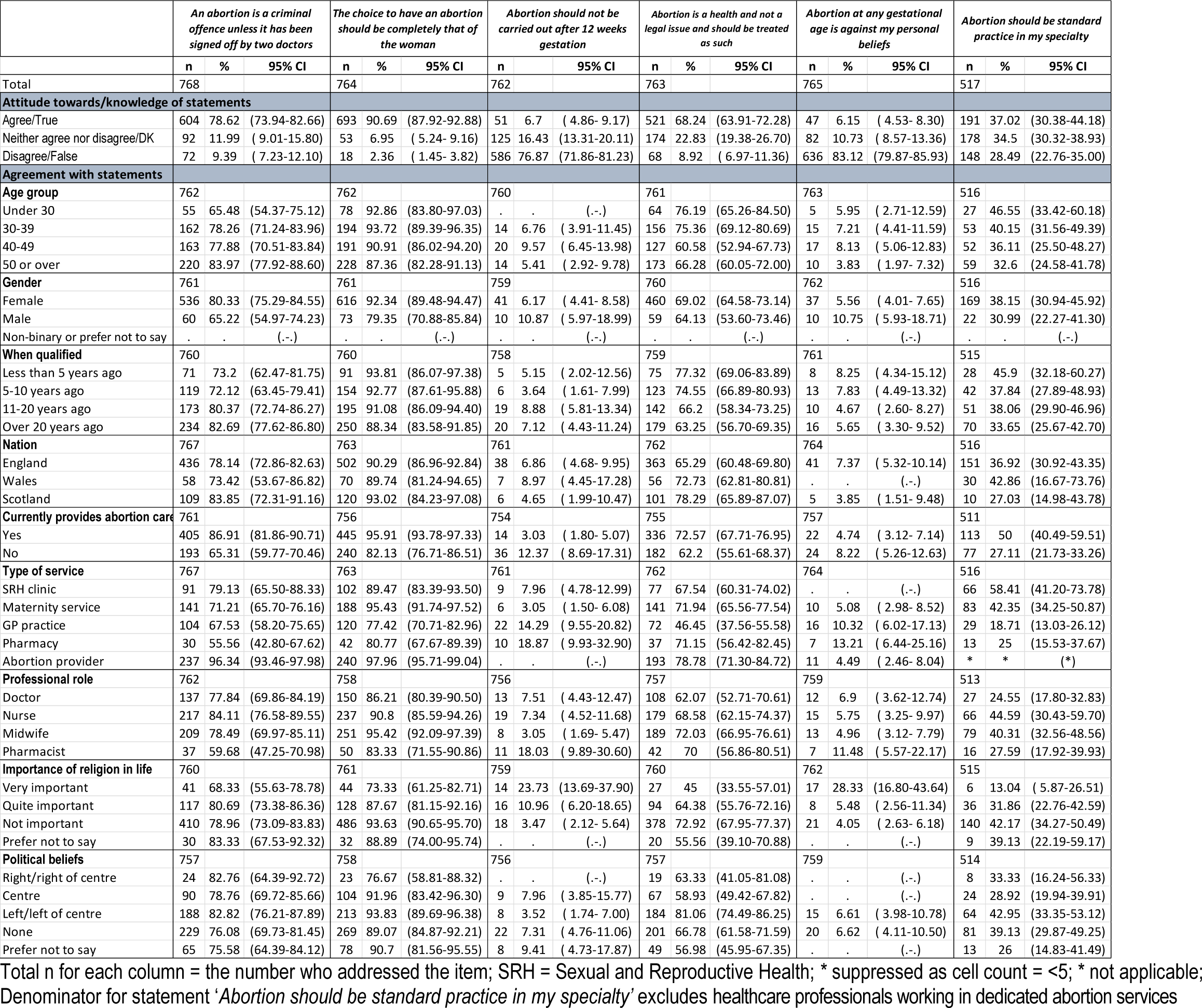
Knowledge of and attitudes towards the regulation of abortion among health care professionals.

**Table 2.** Views of health care professionals on which practitioners should be able to deliver abortion care.

|  | *With training, which practitioners do you feel should be able to deliver the following aspects of care?* |  |  |  |  |  |  |  |  |  |  |  |
| --- | --- | --- | --- | --- | --- | --- | --- | --- | --- | --- | --- | --- |
|  | Doctors |  |  | Nurses |  |  | Midwives |  |  | Pharmacists |  |  |
| **Proportion ticking to endorse** | n | % | 95% CI | n | % | 95% CI | n | % | 95% CI | n | % | 95% CI |
| Prescribing abortion medication | 743 | 96.49 | (94.97-97. | 502 | 65.28 | (59.74-70. | 243 | 31.6 | (27.77-35. | 268 | 34.72 | (31.30-38.30) |
| Dispensing abortion medication | 661 | 85.83 | (82.88-88. | 661 | 85.83 | (82.61-88. | 431 | 56.05 | (50.32-61. | 491 | 63.72 | (60.31-67.00) |
| Carrying out surgical abortion <14 weeks | 730 | 94.8 | (93.08-96. | 314 | 40.7 | (36.78-44. | 104 | 13.52 | (11.07-16. | 18 | 2.34 | ( 1.51- 3.61) |
| Carrying out surgical abortion >=14 weeks | 715 | 92.85 | (90.86-94. | 247 | 31.99 | (28.59-35. | 86 | 11.18 | ( 8.75-14.1 | 16 | 2.08 | ( 1.26- 3.42) |

Note: For doctors and nurses, the proportions represent views on working in any one setting

### Analysis

Survey data were analysed in Stata 17,^20^ taking account of clustering within health service sites. We present descriptive analyses (estimated percentages and 95% confidence intervals) of the knowledge and attitude measures by respondent characteristics. Analysis of data on views on the regulation of abortion included the total sample of healthcare professionals. Analysis of data on views on whether it should be provided in respondents’ own healthcare services excluded providers in dedicated abortion services.

Free text comments relevant to the survey responses were selected for thematic analysis with the aim of better understanding survey responses. Textual data were independently analysed thematically by KW and RF using the Framework method^21,^.

### Patient and public involvement

A Patient and Public Involvement panel was established for the study to help inform study design and recommendations.

## Results

Of 1,370 questionnaires distributed to eligible healthcare professionals, 771 (56.3%) were returned from 147 health service sites. Their characteristics are shown in Supplementary Table x.

### Knowledge

Of all healthcare professionals surveyed, 78.6% selected ‘True’ to the statement: ‘*An abortion is a criminal offence unless it has been signed off by a doctor’*; 9.4% selected ‘False’. The proportion providing the correct answer was higher among women (80.3%) than men (65.2%) and increased with years since qualification and with age (Table 1). A third of health care professionals aged under 30 were unaware of this aspect of the law. Correct understanding of the law was more common among those currently providing abortion in any service (86.9%) compared to those not doing so (65.3%) and was near universal among those working in a specialist abortion service (96.3%). It was less common among those in other service types; almost half of those in pharmacies were unaware of this legal requirement.

### Attitudes towards the regulation of abortion

Agreement with the view that abortion was a woman’s choice was high; 90.7% overall saw it as such and only 2.4% disagreed (Table 2). Agreement was higher among women (92.3%) than men (79.4%), among respondents currently providing abortion (95.9%) compared to those not doing so (82.1%) and among those seeing religion as not important in their lives (93.6%) compared with those for whom it was very important (73.3%). Agreement was also higher among healthcare professionals in maternity services (95.4%) and specialist abortion services (98%) compared with those in general practice (77.4%) and pharmacies (80.8%).

Support for the view that abortion was a health, and not a legal issue was less widespread, but was nevertheless a majority opinion with 68.2% overall agreeing and 8.9% disagreeing.

Agreement was lower among those considering religion to be very important in their lives (45%) compared with others and decreased with time lapse since qualification. Important differences by service type were seen only for healthcare professionals working in general practice, fewer than half of whom (46.5%) endorsed this view.

A small minority, 6.2%, of respondents agreed that abortion at any gestational age was against their personal beliefs, 83.1% disagreed. Levels of agreement were marginally higher among men, respondents with right of centre political views, and those employed in general practice and pharmacies (rising slightly above 10% in each case). Among those for whom religion was very important in their lives, it was considerably higher, at 28.3%.

A similarly small minority, 6.7% agreed with the statement that abortion should not be carried out after 12 weeks gestation; 76.9% disagreed. Endorsement of the statement was more common among respondents aged 40 and over and among those for whom religion was very important.

Again, marked differences were seen by service type. The proportion of healthcare professionals who held this view was higher among staff in general practice and reached one in five in pharmacies (18.9%).

### Attitudes towards provision of abortion in different service settings

Of the 517 respondents employed outside of dedicated abortion services, 37% agreed that abortion care should be standard practice in their specialty; 28.5% disagreed. (Table 1) Differences were seen by service type and professional role. Fewer than one in five healthcare professionals in general practice (18.7%) and one in four of those in pharmacies (25%) supported the idea, compared with the majority of those in sexual and reproductive health services (58.4%) and a sizeable minority (42.9%) of those in maternity services. Receptivity to the idea of abortion provision in their service was higher among nurses and midwives than among doctors, across all services.

### Views on provision of abortion care by different practitioners

All respondents were asked for their views on which practitioners, other than designated abortion providers, should be able with training to provide aspects of abortion care. Support for prescription and provision of abortion medication by such practitioners was higher than for surgical methods being carried out by them. (Table 2) Endorsement of the notion that doctors should be able to prescribe and dispense was near unanimous; 96.5% and 85.8% respectively signified approval. For nurses it was slightly lower but still a majority view; nearly two thirds of respondents (65.3%) felt they should be able to prescribe, and 85.8% that they should be able to dispense. Support for midwives and pharmacists prescribing abortion medication was indicated by roughly a third (31.6% and 34.7% respectively) but more than half felt they should be able to dispense abortion medication.

With regard to surgical methods, again there was strong support for doctors carrying out surgical abortion, with 94.8% indicating that they should be able to perform surgery before 14 weeks and 92.9% after this time. Affirmation of surgical methods carried out by nurses was lower, but a sizeable minority of respondents (41%) felt that nurses should be able to carry out surgical abortion before 14 weeks. Support for surgical methods being carried out by midwives was lower, and by pharmacists was negligible.

### Free text comments

26 free text comments added to the questionnaire related to the regulation of abortion and 56 to the appropriateness of abortion provision in different healthcare settings.

Comments relating to the regulation of abortion were universally in favour of relaxing the law. (Table 3) None stated a preference for retaining the current legal restrictions which were seen as outdated and as having adverse consequences for the efficiency and quality of abortion provision. Where this view was qualified, it was with reference to the need for a prior medical consultation. Strong opposition was expressed to the continued need for two doctors to authorise an abortion. It was held that, should the legal requirements remain in place, since abortion was increasingly led by nurses their role should include responsibility for certifying that the grounds for abortion were met.

**Table 3.**
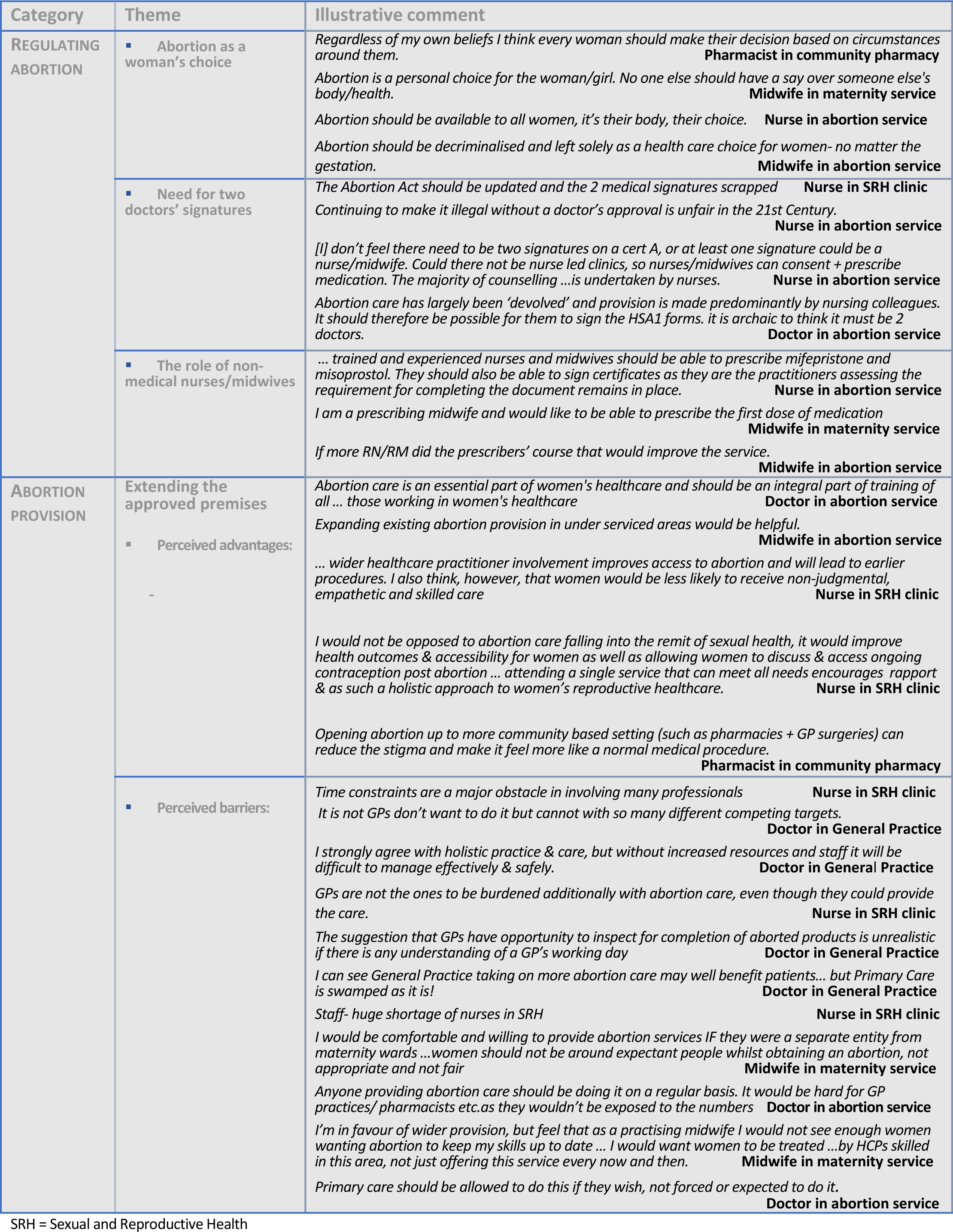
Illustrative examples of free text comments.

There was a consensus that abortion should be nurse-led with the proviso that this should be a provider’s individual choice. Mandating responsibility for abortion provision was considered likely to negatively impact both patients and healthcare professionals. Comments suggested that the lower levels of endorsement for incorporating abortion into standard heath care seen in the survey data may be attributable to recognition of the challenges rather than to disinclination to provide abortion. While benefits of integration were identified, notably reducing stigma surrounding abortion, increasing access and availability of provision, and providing more holistic care, so too were potential costs. These included a possibly detrimental effect on other services, a poorer quality abortion service where skill sets were not adequate and possibly less sensitive treatment of patients. Greater involvement of primary care in abortion provision was seen as currently unfeasible given perceptions that the service was already understaffed and overburdened. Similar, though more muted, reservations were expressed in relation to midwifery with the additional concern that midwives were unlikely to see enough abortion patients to maintain essential skills. Pharmacists were considered useful sources of information but reservations focussed on the availability of private space. By contrast, and amplifying survey responses, clear benefits were identified for abortion provision in community sexual and reproductive health services in terms of continuity of care, provision of a more holistic service and attention from knowledgeable staff who were sympathetic to patients and comfortable with the service they provided. Absorption of the function was seen as conditional on a change to commissioning patterns and addressing resource limitations.

## Discussion

Our data show generally liberal attitudes towards the regulation of abortion among healthcare professionals in Britain. The view that abortion should be completely a woman’s choice is near universal and a clear majority support the idea of abortion being treated as a health as opposed to a legal issue. Fewer than one in 10 saw abortion at any gestational age as contrary to their personal beliefs. Perhaps surprising was the relatively low levels of awareness, especially higher among younger practitioners, of the legal requirement for two registered medical practitioners to authorise an abortion. Free text comments suggested that many practitioners consider the current law to be outdated.

We found strong support in the survey data for nurses being able to prescribe abortion medication and, in free text comments, for them to authorise abortions. Participants’ views on the desirability of incorporating abortion within their existing practice varied markedly by professional role and service, support for the idea being higher among nurses than doctors, and among practitioners in sexual and reproductive health services compared with those in general practice. Free text comments shed light on perceived benefits of integrating abortion into standard health care, including continuity of care, more holistic health care and destigmatising abortion but also the challenges, including constraints of time, staffing levels and resources, especially in general practice.

### Strengths and limitations

Our study provides much needed data on the views of healthcare professionals on abortion regulation. A strength lies in the range of healthcare professionals surveyed, unique in British studies to date, providing opportunities for comparisons to be made between specialities and settings. The study also benefits from use of a stratified cluster sampling strategy, increasing the generalisability of findings. The addition of qualitative data from free text comments provides insights into survey responses. The main limitation stems from the timing of the study. Scheduling fieldwork at the height of the COVID-19 pandemic in Britain and its immediate aftermath may have introduced both participation and reporting bias. Healthcare professionals most actively involved in coping with the pandemic may have declined to participate because of time pressures. The responses of those who did may reflect heightened awareness of the constraints of their workload which may have negatively influenced their propensity to be involved in abortion care and support.

### Interpretation and contextualisation

Comparisons of the findings of this study and those of others are difficult because of differences in questions asked, populations under study, and recency of investigation. We found no other studies exploring knowledge of regulations governing abortion provision. Other studies in Britain have found similarly liberal attitudes among healthcare professionals towards the regulation of abortion. In 2000, a survey showed that four in five GPs considered themselves broadly pro-choice^17^ and in an investigation into the attitudes of medical students in Britain, 73% of medical students did so^16^. Strong associations between attitudes towards abortion and the importance of religion are also seen in other studies^22,16,23,24^. We found no other research which compared attitudes towards abortion by service type or profession. The views of healthcare professionals appear to be generally in line with those of the general public. The most recent British Social Attitudes ^Survey25^ shows 76% support for allowing abortions if the woman does not want the child.

Regarding provision of abortion, our findings are less consistent with those of others. The strong support amongst healthcare professionals in community sexual and reproductive health clinics for abortion provision being standard practice in their service is consistent with evidence that such settings would be sensitive to the needs of abortion patients and better able to meet other sexual and reproductive health needs including ongoing contraception ^26^ ^27^. However, the markedly lower level of enthusiasm for routinely incorporating abortion care and support into general practice sits in tension with recent international evidence suggesting the merits of doing so ^28, 29,30,31^ ^32^. Thus, the recommendation that insights from LMIC countries might translate to the UK setting^28^ needs to be treated with caution. Contextual differences in Britain – notably, the pressures under which NHS is operating and the consequent burden on primary care and other health services – may adjust the cost-benefit ratio and limit knowledge translation from other settings.

### Implications for policy and practice

We anticipate that our findings will be of interest to commissioners and policy makers. First, the data may inform changes to practice within existing legislation. The recognition among healthcare professionals that aspects of the current law are out of step with best practice echoes earlier findings of the RCOG^7^ and the House of Commons Science and Technology Committee^33^ that the requirement for two doctors’ signatures no longer serves a useful purpose. With regard to which practitioners are permitted to provide abortion, the support shown in our study for nurses and midwives to be permitted to authorise abortions and to prescribe abortion medication is consistent with the WHO guideline urging consideration of the wider range of cadres which can safely perform medical and surgical abortion^34^.

At the time of writing, abortions must be performed within NHS hospitals, licensed abortion clinics or other classes of places licensed by the Secretary of State for Health, including the patient’s home. Our findings suggest that consideration might also be given to routinely licensing sexual and reproductive health services to offer abortion care. They suggest the need to proceed with more caution in considering any wholesale extension of abortion care into General Practice. Although a sizeable minority of staff in this specialty would support such a move, offering the potential to increase access - especially in underserved rural areas - the majority would not. While examples can be provided from elsewhere in the world of the potential advantages of integration into general practice, high quality services are unlikely to result where practitioners currently have neither the capacity nor the resources to offer them.

Finally, should there be political will for a legal reform that removes specific criminal prohibitions against abortion, our study suggests that it would encounter strong support amongst healthcare professionals. Our respondents expressed a clear preference for regulating abortion as a matter of health and the individual’s choice, rather than as an issue properly addressed through criminal law.

## Data Availability

All data publicly available from the study are contained in the manuscript

## Acknowledgments

\# The SACHA Study team also includes: Annette Aronsson (Karolinska Institute, Sweden), Paula Baraitser (Kings College London), Sharon Cameron (University of Edinburgh), Caroline Free (LSHTM), Louise Keogh (University of Melbourne, Australia), Patricia A. Lohr (BPAS, UK), Clare Murphy (BPAS), Wendy V. Norman (University of British Columbia, Canada) and Geoff Wong (University of Oxford).

We are grateful to survey participants who gave their time to the study and to service managers who helped identify eligible staff. We would like to thank the North Thames Clinical Research Network for their help and advice; local R&D Departments for assistance in obtaining approvals; and our Advisory Group under the Chairmanship of Dr Jonathan Lord for their constant advice and support.

## Copyright/Licence for Publication

*The Corresponding Author has the right to grant on behalf of all authors and does grant on behalf of all authors, a worldwide licence to the Publishers and its licensees in perpetuity, in all forms, formats and media (whether known now or created in the future), to i) publish, reproduce, distribute, display and store the Contribution, ii) translate the Contribution into other languages, create adaptations, reprints, include within collections and create summaries, extracts and/or, abstracts of the Contribution, iii) create any other derivative work(s) based on the Contribution, iv) to exploit all subsidiary rights in the Contribution, v) the inclusion of electronic links from the Contribution to third party material where-ever it may be located; and, vi) licence any third party to do any or all of the above*.

## Data availability

Data are not publicly available. Restrictions relate to the terms of ethical approval granted.

## Funding

The study was supported by the NIHR [HSDR Project: NIHR129529]. The views expressed are those of the authors and not necessarily those of the NIHR or the Department of Health and Social Care.

## Competing interest

All authors have completed the Unified Competing Interest form and declare: no support from any organisation for the submitted work; no financial relationships with any organisations that might have an interest in the submitted work in the previous three years; and no other relationships or activities that could appear to have influenced the submitted work

## Contributor and Guarantor Information

KW and RF conceptualised the study. RF, MP, JS, KW designed the survey with input from all authors. RF, JS, MP, RM, ML and NS managed ethics/R&D approvals and fieldwork. RS, OM and MP contributed to the analysis. KW drafted the paper with input from SS, RF, RS, MP, RM, ML and JR. All authors approved the final manuscript. The corresponding author KW, guarantor for the work, attests that all listed authors meet authorship criteria and that no others meeting the criteria have been omitted.

## Ethics approval

Approval was obtained from the Research Ethics Committees of the London School of Hygiene & Tropical Medicine (Refs. 22761/26332) and the National Health Service (NHS) (Ref. 21/LO/0236)/ NHS Health Research Authority (IRAS Approval ID 297849).

## Competing interest

All authors have completed the Unified Competing Interest form (available on request from the corresponding author) and declare: no support from any organisation for the submitted work [or describe if any]; no financial relationships with any organisations that might have an interest in the submitted work in the previous three years [or describe if any], no other relationships or activities that could appear to have influenced the submitted work [or describe if any]." Please note: The corresponding author must collect Unified Competing Interest forms from all authors and summarise their declarations as above within the manuscript.

---

**Appendix Table X.**
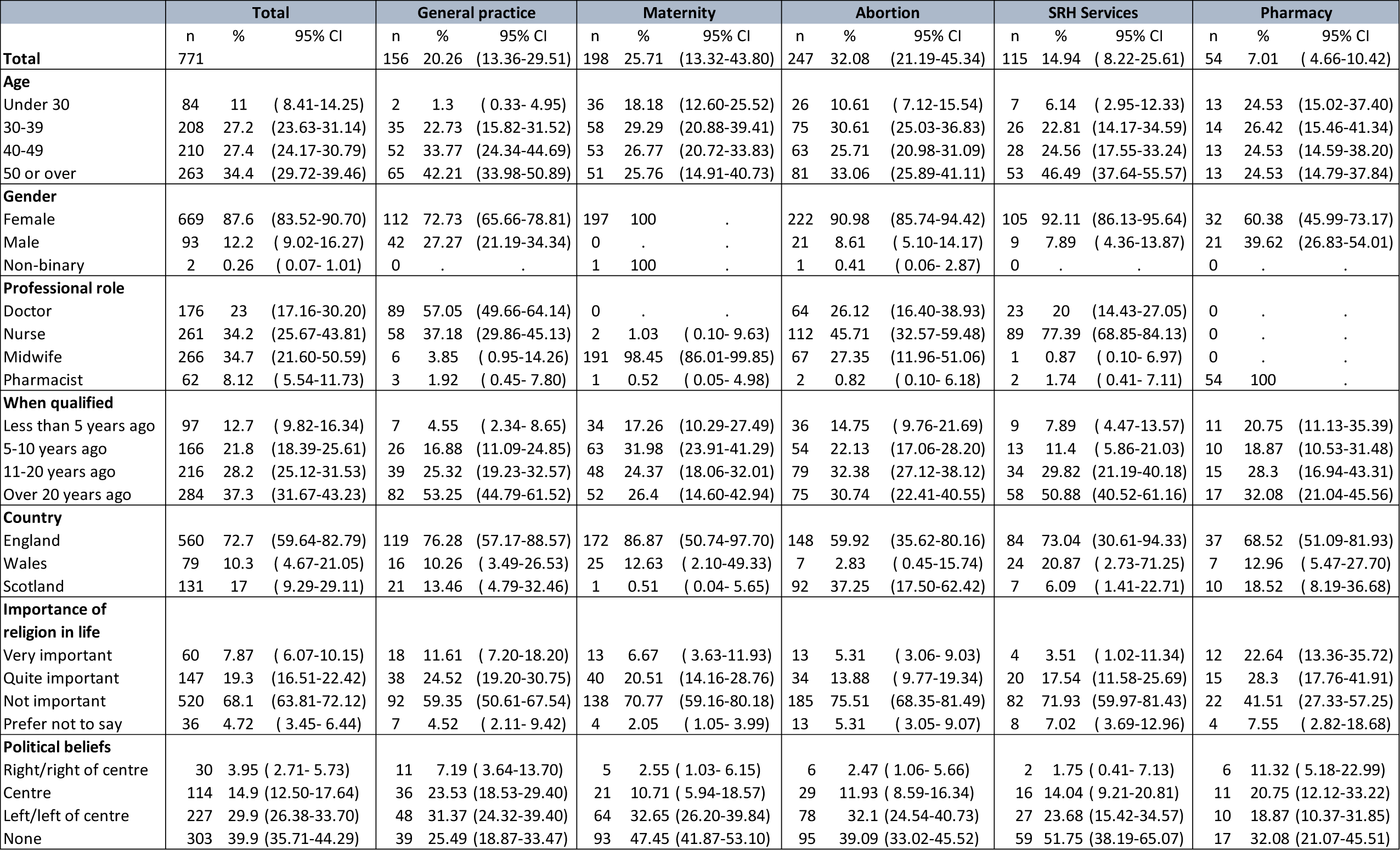
Demographic profile of the sample by service type.

## Footnotes

1 https://www.lshtm.ac.uk/research/centres-projects-groups/sacha

2 https://www.lshtm.ac.uk/research/centres-projects-groups/sacha

## Notes

### Competing Interest Statement

The authors have declared no competing interest.

### Funding Statement

Ethical approval was obtained from the Research Ethics Committees of the London School of Hygiene & Tropical Medicine (Refs. 22761/26332) and the National Health Service (NHS) (Ref. 21/LO/0236)/ NHS Health Research Authority (IRAS Approval ID 297849).

## References

1. Berer M. Reconceptualizing safe abortion and abortion services in the age of abortion pills: A discussion paper. Best Pract Res Clin Obstet Gynaecol. 2020;63:45–55. doi:10.1016/j.bpobgyn.2019.07.012.

2. Sheldon S, Davis G, O’Neill J, Parker C. (2023) The Abortion Act, 1967; A Biography of a UK Law. Cambridge University Press

3. Rough E. Early medical abortion at home during and after the pandemic: 30 Nov 2022. Available: https://commonslibrary.parliament.uk/research-briefings/cbp-9496/ (Accessed 01/06/2023)

4. Centre for Reproductive Rights. The World’s Abortion Laws https://reproductiverights.org/maps/worlds-abortion-laws/ (Accessed 08.01.24)

5. Garré P. The New Abortion Law in Belgium Leads to a Virtually Full Right to the Termination of Pregnancy in the First 12 Weeks. International Journal of Philosophy 2021;9(4):246–252. doi:10.11648/j.ijp.20210904.20 Accessed 10.10. 23

6. Sheldon S, O’Neill J, Parker C, et al. ‘Too Much, too Indigestible, too Fast’? The Decades of Struggle for Abortion Law Reform in Northern Ireland Too Much, too Indigestible, too Fast’? The Decades of Struggle for Abortion Law Reform in Northern Ireland. Modern Law Review 2020; 83(4): 725–928. 10.1111/1468-2230.12521

7. RCOG, Reforming Abortion Law Position Statement. 13 Jun 2023 Reforming Abortion Law Position Statement | RCOG Accessed 13.09.23

8. Women’s health leaders renew calls for the UK Government to decriminalise abortion https://www.rcog.org.uk/news/women-s-health-leaders-renew-calls-for-the-uk-government-to-decriminalise-abortion/;

9. Labour party manifesto. 2019; It’s Time for Real Change. https://labour.org.uk/wp-content/uploads/2019/11/Real-Change-Labour-Manifesto-2019.pdf Accessed 09.11.23

10. UK Parliament; Parliamentary Bills, 2019. https://bills.parliament.uk/bills/2302

11. UN News. WHO issues new guidelines on abortion to help deliver lifesaving care. Accessed 22.09.23

12. Cedaw WDA (2015), Reproductive Rights are Human Rights: A Handbook for National Human Rights Institutions | OHCHR

13. Kishen, M., & Stedman, Y. (2010). The role of Advanced Nurse Practitioners in the availability of abortion services. Best practice & research. Clinical obstetrics & gynaecology 2010;24(5):569–578. 10.1016/j.bpobgyn.2010.02.014

14. Department of Health and Social Care. Abortion Statistics, England and Wales: 2021; 2022 Nov 17; available at https://www.gov.uk/government/statistics/abortion-statistics-for-england-and-wales2021/abortion-statistics-england-and-wales-2021 (accessed 22.09.23).

15. Thornton J. Demand for abortions surges as contraception services shrink and cost of living rises. BMJ 2023 Jan;31;380:237. doi: 10.1136/bmj.p237

16. Gleeson R, Forde E, Bates E, et al. Medical students’ attitudes towards abortion: a UK study. J Med Ethics.2008 Nov;34(11):783–7. 10.1136/jme.2007.023416

17. Francome C, Freeman E. British general practitioners’ attitudes toward abortion. Fam Plann Perspect. 2000 Jul-Aug;32(4):189–91.

18. Savage WD, Francome C. Gynaecologists’ attitude to abortion provision in 2015. J Obstet Gynaecol 2017 Apr;37(3):406–408. doi: 10.1080/01443615.2016.1233950.

19. SACHA study website: https://www.lshtm.ac.uk/research/centres-projects-groups/sacha

20. Stata Statistical Software: Release 18. College Station, TX: StataCorp LLC.

21. Ritchie J, Spencer L, O’Connor W, et al. Qualitative Research Practice: a Guide for Social Science Students and Researchers. 2nd ed. Los Angeles: SAGE; 2013.

22. Lipp A. A review of termination of pregnancy: prevalent health care professional attitudes and ways of influencing them. J Clin Nurs 2008;17(13):1683–8. doi: 10.1111/j.1365-2702.2007.02205.x.

23. Bloomer F, Kavanagh J, Morgan L, et al. Abortion provision in Northern Ireland: the views of health professionals working in obstetrics and gynaecology units. BMJ sexual & reproductive health, 2022;48(1), 35–40. 10.1136/bmjsrh-2020-200959

24. Malikentzou N, Douzenis A, Chatzinikolaou F, et al. Modern bioethical issues: Euthanasia, physician assisted suicide and abortion. Comparative study of attitudes between physicians and law professionals. Psychiatrik. 2022 Mar;28;33(1):49–55. 10.22365/jpsych.2021.043

25. BSA 40: A liberalisation in attitudes? | National Centre for Social Research (natcen.ac.uk) Accessed 10.10.23

26. Michie L, Cameron ST, Glasier A. Abortion care services delivered from a community sexual and reproductive health setting: views of health care professionals. J Fam Plann Reprod Health Care 2013 Oct;39(4):270–5. doi: 10.1136/jfprhc-2012-100563.

27. Cameron ST, Glasier A, Johnstone A. Shifting abortion care from a hospital to a community sexual and reproductive health care setting. J Fam Plann Reprod Health Care. 2016 Apr;42(2):127–32. doi: 10.1136/jfprhc-2015-101177.

27. Maxwell KJ, Hoggart L, Bloomer F, Rowlands S, Purcell C. Normalising abortion: what role can health professionals play? BMJ Sex Reprod Health 2020 Apr; 2;47(1):32–6. 10.1136/bmjsrh-2019-200480

28. Zhou J, Blaylock R, Harris M. Systematic review of early abortion services in low- and middle-income country primary care: potential for reverse innovation and application in the UK context. Global Health 2020 Sep 30;16(1):91. doi: 10.1186/s12992-020-00613.

29. Wolgemuth T, Judge-Golden C, Lane K, et al. Perspectives of internal medicine physicians regarding medication abortion provision in the primary care setting. Contraception 2021 Oct;104(4):420–425. doi: 10.1016/j.contraception.2021.04.012.

30. Subasinghe AK, Deb S, Mazza D. Primary care providers’ knowledge, attitudes and practices of medical abortion: a systematic review. BMJ Sex Reprod Health 2019 Dec; 30:bmjsrh-2019–200487. doi: 10.1136/bmjsrh-2019-200487

31. Schellekens JE, Houtvast CS, Leusink P, et al. Dutch GPs’ views on prescribing mifepristone and misoprostol: a mixed-methods study. Br J Gen Pract 2022 Apr; 29;72(722):e677–83. doi: 10.3399/BJGP.2021.0704

32. Munro S, Guilbert E, Wagner M-S, Wilcox ES, Devane C, Dunn S, et al. Perspectives Among Canadian Physicians on Factors Influencing Implementation of Mifepristone Medical Abortion: A National Qualitative Study. Annals of family medicine. 2020;18(5):413–21.

33. House of Commons, 2007. Scientific developments relating to the Abortion Act, 1967. Scientific and Technology Committee TSO (HC 1045-1). https://publications.parliament.uk/pa/cm200607/cmselect/cmsctech/1045/1045i.pdf Accessed 10.10.23

34. WHO Abortion Care Guideline 2022. https://srhr.org/abortioncare Accessed 09.01.24

